# Cross-setting replication of the associations between maternal health and autism

**DOI:** 10.1101/2025.06.12.25329531

**Authors:** Vahe Khachadourian, Meredith Anderson, Elias Speleman Arildskov, Jakob Grove, Abraham Reichenberg, Sven Sandin, Diana Schendel, Stefan Nygaard Hansen, Lisa A Croen, Magdalena Janecka

## Abstract

**Background:** Autism spectrum disorder (ASD) is a neurodevelopmental condition with early-life origins. Maternal health conditions during pregnancy have been linked to autism risk, but most studies focus on single populations, limiting generalizability. In this study we examined whether associations previously reported in a Danish registry-based study hold in a U.S. cohort.

**Methods:** We analyzed electronic health records of children born between 2010 and 2017 at Kaiser Permanente Northern California (KPNC) and their mothers. Maternal diagnoses were classified as chronic or non-chronic and their potential associations with ASD diagnosis in the child were assessed using Cox proportional hazards models, adjusting for key sociodemographic, healthcare utilization, and comorbid maternal diagnoses. Methods were aligned with the Danish study to ensure comparability.

**Results:** Among 224,353 children in the KPNC cohort, 5,448 (2.4%) were diagnosed with autism. Of the 42 maternal diagnoses significantly associated with autism in Denmark, 38 were evaluable in KPNC, and 18 remained statistically significant after full adjustment. The majority of associations had point estimates consistent with the Danish study, particularly psychiatric and cardiometabolic conditions (e.g., bipolar disorder: HR = 1.62, 95% CI = 1.30–2.02; gestational diabetes: HR = 1.22, 95% CI = 1.12–1.32).

**Conclusions:** Despite demographic and healthcare differences, and some differences in effect magnitudes of maternal health-autism associations, 35 of the 38 associations found in the Danish study replicated, qualitatively (direction of the effect), in the U.S. cohort, suggesting robust cross-setting relevance. Further research is needed to explore underlying mechanisms and potential effect modifiers.

## INTRODUCTION

Autism spectrum disorder (ASD) is a neurodevelopmental condition characterized by differences in social communication and restrictive, repetitive behaviors, often detected in early childhood. For brevity, throughout the article, we use ‘autism’ to refer to a diagnosis of autism spectrum disorder. Given the early-life origins of autism, and the extensive maternal-fetal interface that is established during pregnancy, etiological research has focused on perinatal influences as potential risk factors for autism. Epidemiological studies have identified links between maternal health during pregnancy — including diagnoses of depression^1^, diabetes^2^, immune system disorders^3^, and infections^4^ — and an increased risk of autism in offspring. However, many maternal health conditions have been explored in relation to autism risk individually, often with differing methodologies across the studies. This precludes comparisons of autism risk across different maternal conditions within a study setting, as well as drawing robust conclusions about the extent to which these associations replicate *between* different settings.

Addressing the historically limited scope of within-setting comparisons, our recent registry-based study in Denmark^5^ evaluated a broad spectrum of maternal diagnoses during pregnancy in the context of autism risk in offspring. Of the 236 maternal diagnoses evaluated in that study, 42 were statistically significantly associated with autism in the offspring. While this enabled us to compare the strength of the different associations within the same setting, findings were based on a population that is relatively homogeneous in terms of genetic ancestry, and access to care (Danish). Although Denmark, like other Nordic countries, provides an unparalleled opportunity to identify autism risk factors – due to the population-wide coverage of the health registers, reliable longitudinal person tracking, and universal healthcare – the generalizability of the findings to cohorts with more diverse health care access and demographics, remains unknown.

The extent to which the effects of perinatal risk factors for autism replicate between settings is not well understood, limiting the interpretation of observational associations in a single setting, and reducing the portability of this information for clinical prediction in different settings. Therefore, the current study aims to examine the generalizability of the associations between maternal health conditions during pregnancy and the risk of autism in offspring characterized in the Danish National Register. To this end, we examined the replication of the associations as indicated by the direction and statistical significance of the reported effects - reported in Denmark in a pregnancy cohort with Kaiser Permanente Northern California (KPNC) membership. KPNC has a large membership with diverse sociodemographic characteristics and comprehensive maternal and child health records, enabling close alignment of the analytical methods with those implemented in the Danish study.

## METHODS

The study population included children born within KPNC between January 1, 2010 and December 31, 2017. KPNC is a large, integrated healthcare system that serves over 4.5 million members. The sociodemographic profile of KPNC members closely reflects the local and statewide population of California with the exception of individuals at the highest and lowest ends of the income spectrum who are underrepresented^6,7^.

Children without KPNC enrollment after birth, whose mothers were younger than 13 years or older than 55 years at the time of the child’s birth, or who had missing data on covariates were excluded. Children were also excluded if their mothers did not have KPNC membership for at least 3 months during each year in the 2 years preceding delivery. Children were followed until 1) February 29, 2020, 2) first ASD diagnosis after 1 year of age, 3) disenrollment from KPNC, or 4) death, whichever came first.

### Exposure

Exposures were maternal diagnoses around pregnancy (Table S1), identified using the electronic health records (EHRs) from both inpatient and outpatient settings. We used the Chronic Condition Indicator developed by the Agency for Healthcare Research and Quality^8^ to distinguish between chronic and non-chronic diagnoses. For non-chronic diagnoses, the detection period was the 12 months before childbirth, whereas for chronic diagnoses, the detection period was 48 months before childbirth. The longer detection period for chronic diagnoses reflects the assumption that these conditions are permanent, and their onset cannot be precisely dated using EHR data. A wider detection window enhances sensitivity for capturing maternal chronic conditions that might not be recorded near pregnancy (e.g., well-managed conditions requiring minimal care) but could still impact the fetus. A series of sensitivity analyses assessing the impact of using alternative detection periods is provided elsewhere^5^. Given the study objective, exposures were limited to the 42 diagnoses found to be significantly associated with autism in the study in the Danish population.

### Outcome

Outcome was defined as the first autism diagnosis after 12 months of age recorded in the child EHR and based on the DSM criteria applicable to ASD diagnosis at the time of evaluation (DSM-IV or DSM-5)^9^. Among children in the cohort with autism, 85% were evaluated at a KPNC ASD evaluation center, where a multidisciplinary team employed a standardized protocol, including the Autism Diagnostic Observation Schedule^10^. The remaining 15% received diagnoses from child psychiatrists, developmental-behavioral pediatricians, pediatric neurologists, or general pediatricians.

### Covariates

Covariates included maternal age at delivery, insurance type at delivery (categorized as KPNC insurance vs. Government insurance), race/ethnicity (Asian/Pacific Islander, Black, Hispanic, White, and other/unknown), child’s birth year, and sex, all obtained from KPNC EHRs. Insurance type served as a proxy for socioeconomic status; note this is in contrast to the Danish study, where the SES proxies included maternal income and education, not available in the KPNC records. Additionally, the number of days the mother had any in- or outpatient healthcare encounter in the year preceding the child’s birth provided a proxy for healthcare utilization (including both primary care and specialist encounters; in the Danish study, primary care information was limited to midwife-reported diagnoses during pregnancy).

### Statistical Analysis

We assessed the association of each maternal diagnosis with autism separately in a series of Cox proportional hazard models, incrementally adjusting for the study covariates. The association between each maternal diagnosis and autism in offspring was analyzed adjusting first for child’s sex and birth year, then adding maternal age at delivery, then adding insurance type, and finally adding number of healthcare encounters the mother had during the 12 months preceding the child’s birth.

We used clustering sandwich estimators to account for potential within-family correlation due to the presence of siblings in the dataset. All maternal diagnoses occurring in <5 mothers of children with autism and/or mothers of children without autism were excluded from the analyses to minimize the risk of sparse data bias^11^.

Finally, to adjust for possible comorbidity between diagnoses, all diagnoses evaluated individually were concurrently included in a multi-diagnosis model, adjusting for all covariates. All tests of statistical hypotheses were conducted on the 2-sided 5% level of significance.

All statistical programs were aligned with the original study in the Danish birth cohort^5^.

## RESULTS

We identified 227,195 children born at KPNC from January 1, 2010 to December 31, 2017. After excluding one child (<0.01%, no autism) with unknown sex and 2,841 children (1.2% overall, 59 with autism (1.1%) and 2,782 without autism (1.2%)) whose insurance type at delivery was unknown, the study sample included 224,353 children. The median duration of follow-up was 3.7 years (IQR: 1.3 – 6.4 years). A total of 5,448 (2.4%) children received an ASD diagnosis during the study follow-up period. The median age at diagnosis was 3.0 years (IQR: 2.2 - 4.0 years). The median maternal age at the time of delivery was 31.9 years (IQR: 28.3 – 35.3 years). White children were the largest racial/ethnic group (41.3%) in the sample, followed by Asian/Pacific Islander (26.3%) and Hispanic (23.4%) children. **Table S2** presents the sample characteristics of covariate data by ASD diagnosis status.

### Maternal diagnoses and offspring autism

Of the 42 maternal diagnoses significantly associated with autism in Denmark, 4 diagnoses were observed in fewer than five mothers of children with autism in the KPNC cohort, corresponding to a prevalence of less than 0.1% per diagnosis in KPNC. After excluding these rare diagnoses whose effects could not be reliably estimated in the KPNC sample, 38 diagnoses were evaluated for their association with offspring autism. Prevalence of several of these diagnoses differed substantially in the Danish and Californian cohorts, including over 10-fold differences in postpartum hemorrhage (7.4% in DK vs 0.2% in KPNC) or abnormality of the forces of labor (1% in DK vs. 10.5% in KPNC). Table S3 provides the number of exposed children by autism status.

In the fully adjusted model (Figure 1 and Table S4), 18 diagnoses were statistically significantly associated with autism, all of which with point estimates >1. Apart from multiple gestation, all statistically significant diagnoses had point estimates consistent with those observed in the Danish cohort study (multiple gestation, KPNC: HR = 1.25, 95% CI = 1.11–1.40; DK: HR = 0.85, 95% CI = 0.78-0.93). Maternal diagnoses with a consistent association with autism in the KPNC setting included psychiatric (e.g., bipolar disorder; KPNC: HR = 1.62, 95% = 1.30–2.02; DK: HR = 2.14, 95% CI = 1.43-3.20) and cardiometabolic conditions (e.g., diabetes mellitus in pregnancy; KPNC: HR = 1.22, 95% CI = 1.12–1.32, DK: HR = 1.23, 95% CI = 1.12-1.36; primary hypertension; KPNC: HR = 1.20, 95% = 1.06–1.36; DK: HR = 1.34, 95% CI = 1.08-1.67).

**Figure 1.**
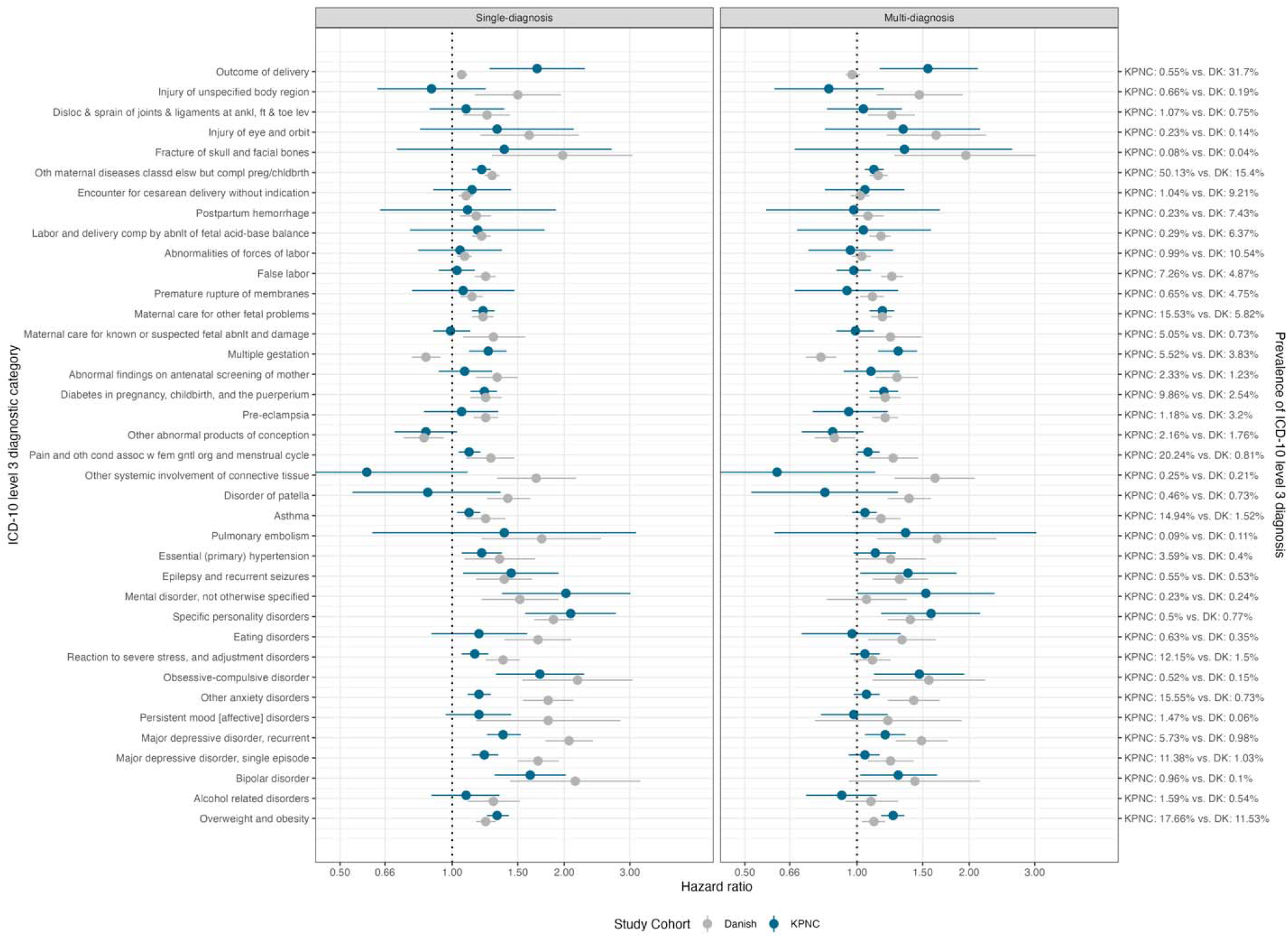
Associations between ICD-10 level 3 maternal diagnoses and offspring autism from single- and multiple-diagnosis models in KPNC and Danish cohorts. All estimates and their confidence interval are from the fully adjusted model accounting for maternal age at delivery, child’s birth year, sex and the measures of SES (insurance type at delivery; race/ethnicity; number of days the mother had any healthcare encounter in the year preceding the child’s birth); NB. In Denmark, the measures of SES included instead maternal income and education, not available in the KPNC records.

Of the 20 diagnoses that were significantly associated with autism in the Danish sample but did not reach statistical significance in the KPNC sample, 16 had point estimates that were qualitatively consistent with those in the Danish study (HR > 1 or HR < 1). For the 4 diagnoses with point estimates in the opposite direction in the Danish and KPNC samples, confidence intervals of the estimated effect were overlapping for all but one diagnosis (Other systemic involvement of connective tissue, DK: HR = 1.68, 95%CI =1.32–2.15; KPNC: HR = 0.59, 95%CI =0.32–1.10; NB. this diagnosis was very rare in KPNC, with only 10 diagnoses in pregnancies leading to an ASD diagnosis in the child). Figure 1 provides a comprehensive comparison of the results from the KPNC and Danish cohorts in the fully adjusted models; a comparison of the intermediate models is presented in Table S4.

Finally, in the multi-diagnosis model, which adjusted for covariates and all 38 maternal diagnoses identified in the fully adjusted single-diagnosis models, we observed consistency of the results across KPNC and Denmark, similar to those observed in the single-diagnosis models. A summary of all 38 associations in the multi-diagnosis models and the respective covariate coefficients are presented in Figure 1 and Table S5.

## DISCUSSION

We tested the replication of the associations between maternal diagnoses around pregnancy and offspring autism in Denmark and an EHR-based cohort in Northern California, encompassing over 200,000 pregnancies with diverse sociodemographic characteristics. Despite the substantial differences in sociodemographic characteristics and study settings, we observed the same direction of the effect in 35 out of 38 of the associations observed in the Danish study. These results suggest the generalizability of the associations between maternal health and child autism beyond the Danish population.

Our findings from the Danish study supported previously reported associations, e.g., between maternal obesity or obsessive-compulsive disorder and autism. Here, we further extend their relevance by demonstrating generalizability to a population beyond Denmark. While other studies have previously indicated the generalizability of certain associations – for example, the association between gestational diabetes and autism has been reported in ^2^ and ^12^– our study is the first to perform such replication in a fully systematic manner by closely aligning the study methods across the settings.

The associations between maternal diagnoses and autism replicated despite the differences in the demographics, health profiles, and healthcare systems between the U.S. and Denmark (Table S6). For example, the prevalence of multiple maternal diagnoses differed substantially between the two studies, which could be attributable to differences in the health profiles, healthcare systems, and surveillance and diagnostic coding practices^13,14^. While ‘perfect’ cross-setting replication is rarely feasible in epidemiological studies, our results indicate that the reported associations are unlikely to arise due to the factors that differ across the settings.

Importantly, replication of the study findings does not necessarily provide evidence for causality of maternal health in autism etiology, as the replicated associations could be attributable to non-causal mechanisms in both cohorts. In fact, the study in Denmark demonstrated that familial confounding was likely key in most of the observed associations between maternal diagnoses and offspring autism. Results from KPNC suggest that these familial factors may overlap across the settings (as would be the case for genetic factors, for example), however, it remains possible that the potential confounders contributing to the associations differ across the settings.

Our study represents the first systematic and comprehensive replication of the associations between maternal health and autism across diverse settings. While in both settings we used large, population-based samples, our approach has several limitations. First, while the associations were largely consistent and robust despite differences in diagnosis prevalence, healthcare systems, population characteristics and length of the follow-up period, there were numerical differences in the magnitude of some of the associations. For the majority (35) of the 38 diagnoses evaluated in this study, the estimated effects had wide confidence intervals, making it difficult to determine whether the observed differences in the estimated associations are due to a true heterogeneity of the effects in different populations and settings or simply reflect statistical uncertainty in the estimates. Secondly, while the Danish study relied on a national registry, the results from KPNC may not generalize across the US; although the cohort includes individuals from a wide range of racial, ethnic, and socioeconomic backgrounds, mirroring the diversity in the U.S., it may underrepresent the 11% of U.S. population, between ages 18-64, that are uninsured^15^.

In conclusion, our results demonstrated a replication of multiple associations between maternal diagnoses and offspring autism across populations and settings. These results indicate the potential cross-setting relevance of the study results and provide early evidence that these associations are attributable to factors that are beyond differences in cohort characteristics between the settings (e.g., demographic composition). Further studies are needed to characterize broader generalizability of these associations, assess potential effect measure modification (e.g., by maternal age), and identify factors contributing to potential effect heterogeneity across different settings.

## Data Availability

We share summary statistics of all association tests performed in the study. Based on the ethical approvals, we are not allowed to share individual patient data. Analytical code will be made available to others, enabling replication by individuals with an approved access to the data resources used in the study.

## TABLES

**Table S1.**

| Diagnosis | ICD-10 code | ICD-9 code |
| --- | --- | --- |
| Other abnormal products of conception | O02 | 631 |
| Pre-eclampsia | O14 | 642 |
| Diabetes mellitus | O24 | 250 |
| Abnormal findings on antenatal screening of mother | O28 | 796 |
| Multiple gestation | O30 | 651 |
| Maternal care for known or suspected fetal abnormality and damage | O35 | 655 |
| Maternal care for other fetal problems | O36 | 656 |
| Premature rupture of membranes | O42 | 658 |
| False labor | O47 | 644 |
| Abnormalities of forces of labor | O62 | 661 |
| Labor and delivery complicated by abnormality of fetal acid-base balance | O68 | 656 |
| Postpartum hemorrhage | O72 | 666 |
| Encounter for cesarean delivery without indication | O82 | 669 |
| Other maternal diseases classifiable elsewhere but complicating pregnancy, childbirth and the puerperium | O99 | 648 |
| Fracture of skull and facial bones | S02 | 803 |
| Injury of eye and orbit | S05 | 918 |
| Dislocation and sprain of joints and ligaments at ankle, foot and toe level | S93 | 845 |
| Injury of unspecified body region | T14 | 959 |
| Poisoning by, adverse effect of and underdosing of psychotropic drugs, not elsewhere classified | T43 | 969 |
| Outcome of delivery | Z37 | V27 |
| Malignant neoplasm of breast | C50 | 174 |
| Overweight or obesity | E66 | 278 |
| Alcohol related disorders | F10 | 291, 303, 305 |
| Schizophrenia | F20 | 295 |
| Schizotypal disorder | F21 | 301 |
| Bipolar disorder | F31 | 296 |
| Depressive episode | F32 | 296 |
| Major depressive disorder, recurrent | F33 | 296 |
| Persistent mood [affective] disorders | F34 | 296 |
| Other anxiety disorders | F41 | 300 |
| Obsessive-compulsive disorder | F42 | 301 |
| Reaction to severe stress, and adjustment disorders | F43 | 308 |
| Eating disorders | F50 | 307 |
| Specific personality disorders | F60 | 301 |
| Unspecified mental disorder | F99 | 300 |
| Epilepsy and recurrent seizures | G40 | 780 |
| Essential (primary) hypertension | I10 | 401 |
| Pulmonary embolism | I26 | 415 |
| Asthma | J45 | 493 |
| Disorder of patella | M22 | 717 |
| Other systemic involvement of connective tissue | M35 | 710 |
| Pain and other conditions associated with female genital organs and menstrual cycle | N94 | 625 |
| Overweight or obesity | E66 | 278 |

**Table S2.** Demographic characteristics of the analytical sample (mother-child dyads)

| **Variables** | **Diagnosed with autism (n=5,448)** | **No autism diagnosis (n=218,905)** | **Total (N=224,353)** |
| --- | --- | --- | --- |
| **Maternal age at delivery, mean (SD)** | 31.8 (5.6) | 31.1 (5.4) | 31.1 (5.4) |
| **Child's sex, n (%)** |  |  |  |
| Female | 1,103 (20.2%) | 108,545 (49.6%) | 109,648 (48.9%) |
| Male | 4,345 (79.8%) | 110,360 (50.4%) | 114,705 (51.1%) |
| **Child's year of birth, n (%)** |  |  |  |
| 2010 | 620 (11.4%) | 25,146 (11.5%) | 25,766 (11.5%) |
| 2011 | 628 (11.5%) | 25,400 (11.6%) | 26,028 (11.6%) |
| 2012 | 703 (12.9%) | 26,112 (11.9%) | 26,815 (12.0%) |
| 2013 | 741 (13.6%) | 26,089 (11.9%) | 26,830 (12.0%) |
| 2014 | 785 (14.4%) | 27,200 (12.4%) | 27,985 (12.5%) |
| 2015 | 798 (14.6%) | 28,311 (12.9%) | 29,109 (13.0%) |
| 2016 | 734 (13.5%) | 29,881 (13.6%) | 30,615 (13.6%) |
| 2017 | 439 (8.1%) | 30,766 (14.0%) | 31,205 (13.9%) |
| **Maternal number of days with health care visits the year leading up to delivery, n (%)\*** |  |  |  |
| 0-12 | 1112 (20.4%) | 56,723 (25.9%) | 57,835 (25.8%) |
| 13-16 | 1335 (24.5%) | 63,080 (28.8%) | 64,415 (28.7%) |
| 17-21 | 1126 (20.7%) | 46,206 (21.1%) | 47,332 (21.1%) |
| 22+ | 1875 (34.4%) | 52,896 (24.2%) | 54,771 (24.4%) |
| **Race/Ethnicity** |  |  |  |
| Asian/Pacific Islander | 1,693 (31.1%) | 57,300 (26.2%) | 58,993 (26.3%) |
| Black | 493 (9.0%) | 15,004 (6.8%) | 15,497 (6.9%) |
| Hispanic | 1,309 (24.0%) | 51,172 (23.4%) | 52,481 (23.4%) |
| Other/Unknown | 115 (2.1%) | 4,691 (2.1%) | 4,806 (2.1%) |
| White | 1,838 (33.7%) | 90,738 (41.4%) | 92,576 (41.3%) |
| **Insurance Type** |  |  |  |
| Government | 552 (10.1%) | 20,135 (9.2%) | 20,687 (9.2%) |
| KPNC | 4,896 (89.9%) | 198,770 (90.8%) | 203,666 (90.8%) |

\* This also includes primary health diagnoses at KPNC, but not in DK (unless made by a midwife during pregnancy)

**Table S3.** Frequency of maternal diagnosis by autism status in KPNC and Danish cohorts.

| ICD10 | Dx description | KPNC |  | Denmark |  |
| --- | --- | --- | --- | --- | --- |
|  |  | autism | No autism | autism | No autism |
| O02 | Other abnormal products of conception | 115 (2.1%) | 4,730 (2.2%) | 292 (1.59%) | 19,583 (1.76%) |
| O14 | Pre-eclampsia | 82 (1.5%) | 2,566 (1.2%) | 719 (3.91%) | 35,447 (3.18%) |
| O24 | Diabetes in pregnancy, childbirth, and the puerperium | 767 (14%) | 21,364 (9.8%) | 524 (2.85%) | 28,201 (2.53%) |
| O28 | Abnormal findings on antenatal screening of mother | 141 (2.6%) | 5,079 (2.3%) | 254 (1.38%) | 13,618 (1.22%) |
| O30 | Multiple gestation | 439 (8.1%) | 11,936 (5.5%) | 608 (3.31%) | 42,738 (3.84%) |
| O35 | Maternal care for known or suspected fetal abnlty and damage | 333 (6.1%) | 10,988 (5.0%) | 116 (0.63%) | 8,162 (0.73%) |
| O36 | Maternal care for other fetal problems | 1,032 (19%) | 33,821 (15%) | 1,099 (5.98%) | 64,811 (5.82%) |
| O42 | Premature rupture of membranes | 41 (0.8%) | 1,414 (0.6%) | 874 (4.76%) | 52,849 (4.75%) |
| O47 | False labor | 390 (7.2%) | 15,898 (7.3%) | 1,078 (5.87%) | 54,036 (4.85%) |
| O62 | Abnormalities of forces of labor | 62 (1.1%) | 2,151 (1.0%) | 2,233 (12.15%) | 117,106 (10.52%) |
| O68 | Labor and delivery comp by abnlty of fetal acid-base balance | 23 (0.4%) | 628 (0.3%) | 1,291 (7.03%) | 70,816 (6.36%) |
| O72 | Postpartum hemorrhage | 13 (0.2%) | 497 (0.2%) | 539 (2.93%) | 83,559 (7.5%) |
| O82 | Encounter for cesarean delivery without indication | 68 (1.2%) | 2,267 (1.0%) | 2,057 (11.2%) | 102,180 (9.18%) |
| O99 | Oth maternal diseases classd elsw but compl preg/chldbrth | 3,076 (56%) | 109,394 (50%) | 2,391 (13.01%) | 171,945 (15.44%) |
| S02 | Fracture of skull and facial bones | 9 (0.2%) | 180 (<0.1%) | 21 (0.11%) | 485 (0.04%) |
| S05 | Injury of eye and orbit | 17 (0.3%) | 508 (0.2%) | 43 (0.23%) | 1,488 (0.13%) |
| S93 | Disloc & sprain of joints & ligaments at ankl, ft & toe lev | 73 (1.3%) | 2,326 (1.1%) | 212 (1.15%) | 8,227 (0.74%) |
| T14 | Injury of unspecified body region | 35 (0.6%) | 1,450 (0.7%) | 60 (0.33%) | 2,077 (0.19%) |
| T43 | Psychotropic drugs, not elsewhere classified | 4 (<0.1%) | 163 (<0.1%) | 11 (0.06%) | 187 (0.02%) |
| Z37 | Outcome of delivery | 50 (0.9%) | 1,175 (0.5%) | 6,523 (35.5%) | 352,301 (31.64%) |
| C50 | Malignant neoplasm of breast | 4 (<0.1%) | 130 (<0.1%) | 19 (0.1%) | 530 (0.05%) |
| E66 | Overweight and obesity | 1,240 (23%) | 38,379 (18%) | 1,445 (7.86%) | 129,044 (11.59%) |
| F10 | Alcohol related disorders | 97 (1.8%) | 3,478 (1.6%) | 159 (0.87%) | 5,915 (0.53%) |
| F20 | Schizophrenia | 2 (<0.1%) | 89 (<0.1%) | 37 (0.2%) | 1,216 (0.11%) |
| F21 | Schizotypal disorder | 0 (0%) | 5 (<0.1%) | 22 (0.12%) | 430 (0.04%) |
| F31 | Bipolar disorder | 99 (1.8%) | 2,064 (0.9%) | 24 (0.13%) | 1,054 (0.09%) |
| F32 | Major depressive disorder, single episode | 797 (15%) | 24,724 (11%) | 256 (1.39%) | 11,361 (1.02%) |
| F33 | Major depressive disorder, recurrent | 458 (8.4%) | 12,387 (5.7%) | 204 (1.11%) | 10,847 (0.97%) |
| F34 | Persistent mood [affective] disorders | 106 (1.9%) | 3,184 (1.5%) | 19 (0.1%) | 665 (0.06%) |
| F41 | Other anxiety disorders | 1,040 (19%) | 33,845 (15%) | 166 (0.9%) | 8,101 (0.73%) |
| F42 | Obsessive-compulsive disorder | 59 (1.1%) | 1,107 (0.5%) | 37 (0.2%) | 1,651 (0.15%) |
| F43 | Reaction to severe stress, and adjustment disorders | 802 (15%) | 26,456 (12%) | 380 (2.07%) | 16,599 (1.49%) |
| F50 | Eating disorders | 48 (0.9%) | 1,367 (0.6%) | 96 (0.52%) | 3,890 (0.35%) |
| F60 | Specific personality disorders | 68 (1.2%) | 1,062 (0.5%) | 289 (1.57%) | 8,454 (0.76%) |
| F99 | Mental disorder, not otherwise specified | 30 (0.6%) | 479 (0.2%) | 70 (0.38%) | 2,660 (0.24%) |
| G40 | Epilepsy and recurrent seizures | 47 (0.9%) | 1,196 (0.5%) | 144 (0.78%) | 5,838 (0.52%) |
| I10 | Essential (primary) hypertension | 296 (5.4%) | 7,756 (3.5%) | 85 (0.46%) | 4,445 (0.4%) |
| I26 | Pulmonary embolism | 8 (0.1%) | 184 (<0.1%) | 28 (0.15%) | 1,234 (0.11%) |
| J45 | Asthma | 953 (17%) | 32,576 (15%) | 295 (1.61%) | 16,924 (1.52%) |
| M22 | Disorder of patella | 21 (0.4%) | 1,001 (0.5%) | 225 (1.22%) | 8,068 (0.72%) |
| M35 | Other systemic involvement of connective tissue | 10 (0.2%) | 560 (0.3%) | 66 (0.36%) | 2,343 (0.21%) |
| N94 | Pain and oth cond assoc w fem gntl org and menstrual cycle | 1,287 (24%) | 44,122 (20%) | 188 (1.02%) | 9024 (0.81%) |

**Table S4.**
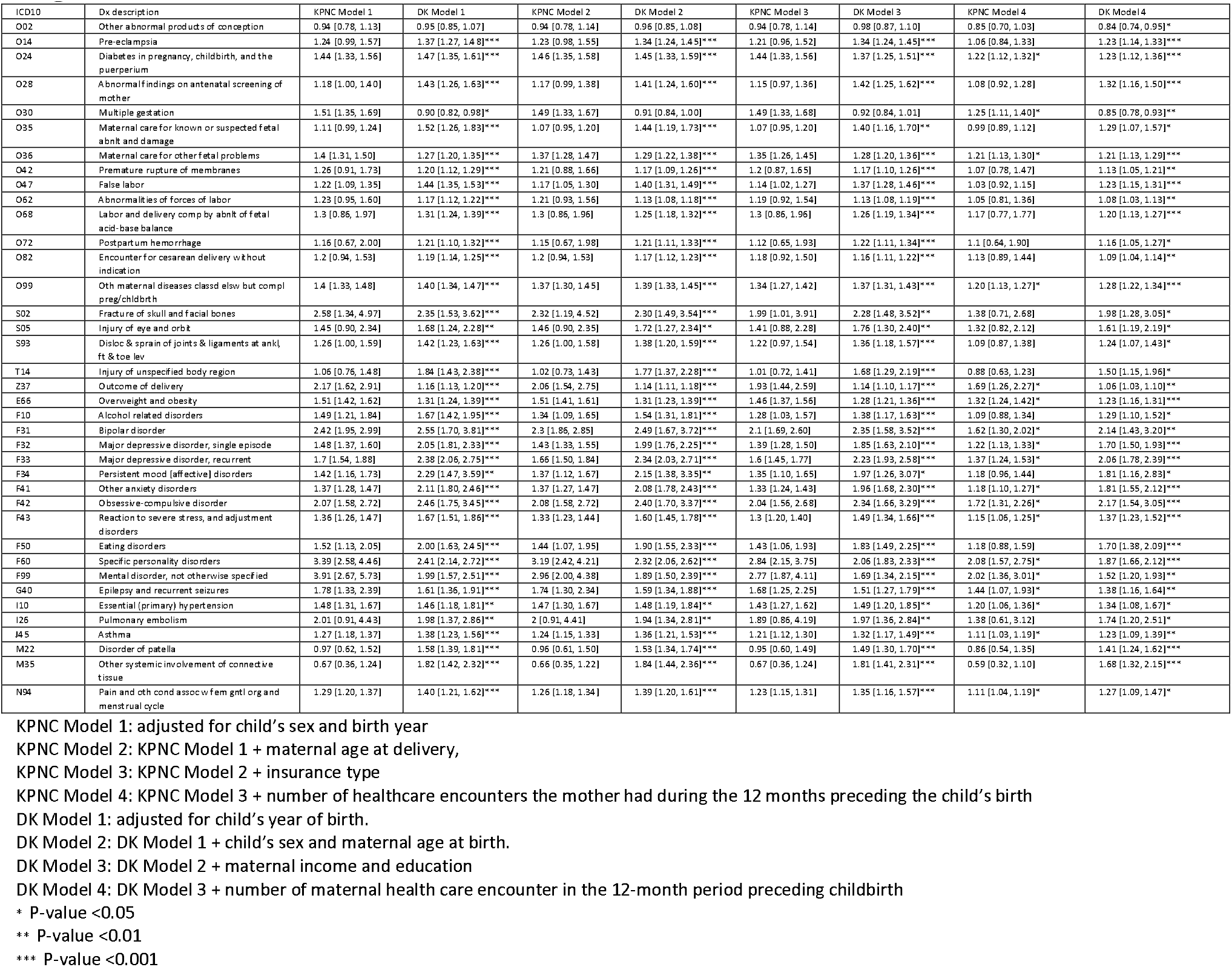
Associations between ICD-10 level 3 maternal diagnoses and offspring autism from single-diagnosis models in KPNC and Danish cohorts.

**Table S5.** Associations between ICD-10 level 3 maternal diagnoses and offspring autism from multi-diagnosis models in KPNC and Danish cohorts.

| ICD10 | DX | KPNC Model 5 | DK Model 5 |
| --- | --- | --- | --- |
| O02 | Other abnormal products of conception | 0.86 [0.71, 1.04] | 0.87 [0.77, 0.99]* |
| O14 | Pre-eclampsia | 0.95 [0.76, 1.21] | 1.19 [1.10, 1.29]*** |
| O24 | Diabetes in pregnancy, childbirth, and the puerperium | 1.18 [1.08, 1.29] | 1.19 [1.08, 1.31]*** |
| O28 | Abnormal findings on antenatal screening of mother | 1.09 [0.92, 1.30] | 1.28 [1.12, 1.46]*** |
| O30 | Multiple gestation | 1.29 [1.14, 1.45] | 0.80 [0.73, 0.88]*** |
| O35 | Maternal care for known or suspected fetal abnlty and damage | 0.99 [0.88, 1.11] | 1.23 [1.01, 1.49]* |
| O36 | Maternal care for other fetal problems | 1.17 [1.08, 1.26] | 1.17 [1.09, 1.24]*** |
| O42 | Premature rupture of membranes | 0.94 [0.68, 1.29] | 1.10 [1.02, 1.18]** |
| O47 | False labor | 0.98 [0.88, 1.09] | 1.24 [1.16, 1.33]*** |
| O62 | Abnormalities of forces of labor | 0.96 [0.74, 1.25] | 1.03 [0.97, 1.09] |
| O68 | Labor and delivery comp by abnlty of fetal acid-base balance | 1.04 [0.69, 1.58] | 1.16 [1.08, 1.23]*** |
| O72 | Postpartum hemorrhage | 0.98 [0.57, 1.67] | 1.07 [0.97, 1.18] |
| O82 | Encounter for cesarean delivery without indication | 1.05 [0.82, 1.34] | 1.02 [0.96, 1.08] |
| O99 | Oth maternal diseases classd elsw but compl preg/chlbrth | 1.11 [1.05, 1.18] | 1.14 [1.08, 1.21]*** |
| S02 | Fracture of skull and facial bones | 1.34 [0.68, 2.61] | 1.96 [1.26, 3.02]** |
| S05 | Injury of eye and orbit | 1.33 [0.82, 2.14] | 1.63 [1.20, 2.22]** |
| S93 | Disloc & sprain of joints & ligaments at ankle, foot & toe lev | 1.04 [0.83, 1.32] | 1.24 [1.07, 1.43]** |
| T14 | Injury of unspecified body region | 0.84 [0.60, 1.18] | 1.47 [1.13, 1.92]** |
| Z37 | Outcome of delivery | 1.55 [1.15, 2.11] | 0.97 [0.93, 1.02] |
| E66 | Overweight and obesity | 1.25 [1.16, 1.34] | 1.11 [1.03, 1.19]** |
| F10 | Alcohol related disorders | 0.91 [0.73, 1.13] | 1.09 [0.93, 1.29] |
| F31 | Bipolar disorder | 1.29 [1.02, 1.64] | 1.43 [0.95, 2.14] |
| F32 | Major depressive disorder, single episode | 1.05 [0.95, 1.15] | 1.23 [1.07, 1.42]** |
| F33 | Major depressive disorder, recurrent | 1.19 [1.05, 1.35] | 1.49 [1.27, 1.75]*** |
| F34 | Persistent mood [affective] disorders | 0.98 [0.80, 1.21] | 1.21 [0.77, 1.91] |
| F41 | Other anxiety disorders | 1.06 [0.98, 1.15] | 1.42 [1.21, 1.67]*** |
| F42 | Obsessive-compulsive disorder | 1.47 [1.11, 1.94] | 1.56 [1.10, 2.21]* |
| F43 | Reaction to severe stress, and adjustment disorders | 1.05 [0.96, 1.15] | 1.10 [0.98, 1.23] |
| F50 | Eating disorders | 0.97 [0.71, 1.31] | 1.32 [1.07, 1.63]* |
| F60 | Specific personality disorders | 1.58 [1.16, 2.14] | 1.39 [1.21, 1.60]*** |
| F99 | Mental disorder, not otherwise specified | 1.53 [1.00, 2.34] | 1.06 [0.83, 1.36] |
| G40 | Epilepsy and recurrent seizures | 1.37 [1.02, 1.85] | 1.30 [1.10, 1.55]** |
| I10 | Essential (primary) hypertension | 1.12 [0.98, 1.27] | 1.23 [0.99, 1.53] |
| I26 | Pulmonary embolism | 1.35 [0.60, 3.03] | 1.64 [1.13, 2.37]** |
| J45 | Asthma | 1.05 [0.97, 1.13] | 1.16 [1.03, 1.31]* |
| M22 | Disorder of patella | 0.82 [0.52, 1.29] | 1.38 [1.21, 1.58]*** |
| M35 | Other systemic involvement of connective tissue | 0.61 [0.33, 1.12] | 1.62 [1.26, 2.07]*** |
| N94 | Pain and oth cond assoc w fem gntl org and menstrual cycle | 1.07 [1.00, 1.15] | 1.25 [1.08, 1.46]** |

KPNC Model 5: adjusted for child's sex and birth year, maternal age at delivery, insurance type, number of healthcare encounters the mother had during the 12 months preceding the child's birth, and all diagnoses in this Table S5
\* P-value <0.05
\*\* P-value <0.01
\*\*\* P-value <0.001

**Table S6.** Demographic characteristics of the analytical sample (mother-child dyads)

| Variables | Diagnosed with autism<br>(n=18,374) | No autism diagnosis<br>(n=1,113,525) | Total<br>(N=1,131,899) |
| --- | --- | --- | --- |
| Maternal age at delivery, mean (SD) | 30.3 (5.1) | 30.6 (4.9) | 30.6 (4.9) |
| Child's sex, n (%) |  |  |  |
| Female | 4,509 (24.5%) | 546,916 (49.1%) | 551,425 (48.7%) |
| Male | 13,865 (75.5%) | 566,609 (50.9%) | 580,474 (51.3%) |
| Child's year of birth, n (%) |  |  |  |
| 1998 | 1,750 (9.5%) | 64,381 (5.8%) | 66,131 (5.8%) |
| 1999 | 1,875 (10.2%) | 64,365 (5.8%) | 66,240 (5.9%) |
| 2000 | 1,936 (10.5%) | 65,175 (5.9%) | 67,111 (5.9%) |
| 2001 | 1,754 (9.5%) | 63,695 (5.7%) | 65,449 (5.8%) |
| 2002 | 1,631 (8.9%) | 62,484 (5.6%) | 64,115 (5.7%) |
| 2003 | 1,526 (8.3%) | 63,133 (5.7%) | 64,659 (5.7%) |
| 2004 | 1,333 (7.3%) | 63,358 (5.7%) | 64,691 (5.7%) |
| 2005 | 1,237 (6.7%) | 63,123 (5.7%) | 64,360 (5.7%) |
| 2006 | 1,147 (6.2%) | 63,891 (5.7%) | 65,038 (5.7%) |
| 2007 | 1,010 (5.5%) | 63,200 (5.7%) | 64,210 (5.7%) |
| 2008 | 838 (4.6%) | 64,248 (5.8%) | 65,086 (5.8%) |
| 2009 | 732 (4.0%) | 62,203 (5.6%) | 62,935 (5.6%) |
| 2010 | 648 (3.5%) | 62,879 (5.6%) | 63,527 (5.6%) |
| 2011 | 460 (2.5%) | 58,662 (5.3%) | 59,122 (5.2%) |
| 2012 | 289 (1.6%) | 57,762 (5.2%) | 58,051 (5.1%) |
| 2013 | 142 (0.8%) | 55,814 (5.0%) | 55,956 (4.9%) |
| 2014 | 53 (0.3%) | 56,874 (5.1%) | 56,927 (5.0%) |
| 2015 | 13 (0.1%) | 58,278 (5.2%) | 58,291 (5.1%) |
| Maternal income in DKK the year before delivery, mean (SD) | 203,452 (110107) | 235,762 (214650) | 235,237 (213400) |
| Missing, n (%) | 103 (0.6%) | 7,844 (0.7%) | 7,947 (0.7%) |
| Paternal income in DKK the year before delivery, mean (SD) | 276,857 (196233) | 316,626 (350731) | 315,980 (348806) |
| Missing, n (%) | 341 (1.9%) | 20,864 (1.9%) | 21,205 (1.9%) |
| Maternal education the year before delivery, n (%) |  |  |  |
| Primary and lower secondary level | 4,828 (26.3%) | 211,468 (19.0%) | 216,296 (19.1%) |
| Upper secondary level and secondary vocational education | 7,844 (42.7%) | 447,248 (40.2%) | 455,092 (40.2%) |
| Short cycle tertiary, bachelor or equivalent | 3,865 (21.0%) | 292,433 (26.3%) | 296,298 (26.2%) |
| Master, doctoral or equivalent | 1,217 (6.6%) | 106,567 (9.6%) | 107,784 (9.5%) |
| Missing, n (%) | 620 (3.4%) | 55,809 (5.0%) | 56,429 (5.0%) |

